# Comparison of clinical metagenomics with 16S rDNA Sanger sequencing for the bacteriological diagnosis of culture-negative samples

**DOI:** 10.1101/2024.06.18.24309080

**Authors:** Camille d’Humières, Skerdi Haviari, Marie Petitjean, Laurène Deconinck, Signara Gueye, Nathan Peiffer-Smadja, Lynda Chalal, Naima Beldjoudi, Geoffrey Rossi, Yann Nguyen, Charles Burdet, Ségolène Perrineau, Diane Le Pluart, Roza Rahli, Michael Thy, Piotr Szychowiak, Xavier Lescure, Véronique Leflon-Guibout, Victoire de Lastours, Etienne Ruppé

## Abstract

**Background:** Currently, diagnosis of bacterial infections is based on culture, possibly followed by the amplification and sequencing (Sanger method) of the 16S rDNA - encoding gene when cultures are negative. Clinical metagenomics (CMg), i.e. the sequencing of a sample’s entire nucleic acids, may allow for the identification of bacteria not detected by conventional methods. Here, we tested the performance of CMg compared to 16S rDNA sequencing (Sanger) in 50 patients with suspected bacterial infection but negative cultures.

**Methods:** This is a prospective cohort study. Fifty patients (73 samples) with negative culture and a 16S rDNA sequencing demand (Sanger) were recruited from two sites. On the same samples, CMg was also performed and compared to 16S rDNA Sanger sequencing. Bacteria were identified using MetaPhlAn4.

**Results:** Among the 73 samples, 20 (27.4%, 17 patients) had a clinically significant 16S rDNA Sanger sequencing result (used for patient management) while 11 (15.1%, 9 patients) were considered contaminants. At the patient level, the sensitivity of CMg was 70.1% (12/17) compared to 16S rDNA. In samples negative for 16S rDNA Sanger sequencing (n=53), CMg identified clinically-relevant bacteria in 10 samples (18.9%, 10 patients) with 14 additional bacteria.

**Conclusions:** CMg was not 100% sensitive when compared to 16S, supporting that it may not be a suitable replacement. However, CMg did find additional bacteria in samples negative for 16S rDNA Sanger. CMg could therefore be positioned as a complementary to 16S rDNA Sanger sequencing.

## Introduction

The diagnosis of bacterial infections relies on culturing clinical samples (e.g. urine, sputum, blood and wounds). However, this method is restricted by its ability to only identify bacteria that can be cultivated under normal laboratory conditions. There is a need for a more comprehensive approach that goes beyond these limitations. Indeed, a large number of potentially pathogenic bacteria (such as strict anaerobic bacteria and intracellular bacteria) are difficult to culture under routine laboratory conditions. In addition, antibiotics taken by the patient prior to sample collection can hinder the growth of bacteria.

To address the limitations of traditional culture methods, scientists have developed “culture-independent” techniques. One such method is 16S ribosomal RNA (rRNA) gene sequencing using Sanger sequencing. This technique involves amplifying and sequencing a highly conserved gene (16S rRNA) found in bacteria. The specific sequence of this gene allows for identification of the bacterial species present in a sample. 16S rDNA Sanger sequencing is particularly useful when cultures are negative but there is a strong suspicion of bacterial infection, such as in cases of suspected infective endocarditis, osteoarticular infections, or meningitis (1). Although 16S rDNA Sanger sequencing is a technique used routinely in clinical microbiology laboratories, it has several limitations: It can be difficult to interpret in cases of polymicrobial infections and does not provide information on potential acquired resistance of the identified bacteria.

Clinical metagenomics (CMg), in contrast, involves sequencing DNA from a sample and then applying bioinformatics tools to extract clinical information. This includes identifying pathogens and predicting their susceptibility to antimicrobials (2). The metagenome of a sample, containing all the genomes of present organisms, holds immense potential for bacteriological diagnosis. It allows for a direct identification of pathogenic bacteria and genes/mutations associated with antibiotic resistance. This concept of CMg emerged alongside the development of new DNA sequencing technologies (“next-generation sequencing” or NGS) in the mid-2000s. CMg has since been applied to a wide range of clinical scenarios (3).

While several studies have compared 16S rDNA Sanger sequencing to 16S rDNA NGS sequencing (4–6), only a few have compared 16S rDNA Sanger sequencing to CMg (using shotgun metagenomics). In a study by Lamoureux *et al*., the results obtained from 16S rDNA Sanger sequencing were compared to those from clinical metagenomics (CMg) in 67 samples (from 64 patients) (7). They reported a high sensitivity of CMg (performed by Illumina-based shotgun metagenomics) with 96.2% at the genus level and after clinical adjudication. In a previous study from our group, we compared the results obtained by CMg to those obtained by conventional methods on bone and joint infection samples (8). We found that the sensitivity of CMg was 100% when compared to 16S rDNA Sanger sequencing. Last, Gu *et al*. compared CMg to various methods including 16S rDNA sequencing in 87 body fluids samples (9). In 8 cases, both methods were performed. CMg and 16S rDNA sequencing yielded consistent results in 6 cases and discordant results in 2.

Here, we aimed to compare the results of clinical metagenomics to those obtained by 16S rDNA Sanger sequencing performed in routine, using a human DNA depletion step on fresh samples during DNA extraction. In addition, we aimed at identifying the clinical, biological and bioinformatic factors associated with CMg positivity.

## Methods

### Population

Meta-16S (“Benefits of metagenomic sequencing for bacteriological diagnosis: comparison with 16S PCR”) is a prospective cohort study conducted between October 2020 and November 2021 at two tertiary-care teaching hospitals: Bichat-Claude Bernard and Beaujon (both located in the Paris area). The study included adult patients suspected of having a bacterial infection who underwent both bacteriological sample collection and a request for 16S rDNA Sanger sequencing on those samples. Patients were excluded if they refused to participate in the study or if the sample quantity was insufficient (<500 µL) for both conventional analyses and DNA extraction. Upon obtaining patient consent, a clinical case report form was completed. The project was approved by the ‘Comité de Protection des Personnes Sud-Ouest et Outre Mer 1’ on October 25, 2021 (2019-A01117-50 – CPP 1-19-077 / SI 5294). Positive results from 16S rDNA Sanger sequencing were considered significant only if the identified bacterium was likely responsible for the infection and was targeted by the patient’s antibiotic regimen.

### DNA manipulations

DNA was extracted from fresh samples using the Ultra-Deep Microbiome Prep (Molzym, Bremen, Germany), which includes a human DNA depletion step. Libraries were prepared using the Nextera XT (Illumina, San Diego, CA), and samples were sequenced in batches on an Illumina NextSeq 500 platform. Quantitative PCR (qPCR) targeting the 16S rRNA gene was performed, using *Escherichia coli* K12 genomic DNA as a calibrator. Each run included a positive control (mock community), a negative biological control (BC, e.g: culture-negative cardiac valves with no suspicion of bacterial infection), and a non-template control (NTC, sterile water). To optimize library preparation, amplification cycles were adjusted using biological negative controls (n=13) and NTC (n=10).

The 16S rDNA Sanger sequencing method employed a semi-automated approach using the UMD-SelectNA CE-IVD kit (Molzym GmbH, Bremen, Germany) for DNA extraction, real-time PCR, and sequencing, as per the manufacturer’s instructions. A DNA control fragment provided by the manufacturer was added to all samples before extraction to evaluate both DNA extraction efficiency and the presence of potential PCR inhibitors. Negative extraction and PCR controls were also included. Amplification of the 16S rRNA - encoding gene targeted the hypervariable region V3-V4 of the bacterial gene. For positive samples, sequencing utilized primers included in the UMD-SelectNA CE-IVD kit, with one pair specific for Gram-positive and another for Gram-negative bacteria. Sequencing was performed on an ABI 3130xl instrument (Thermo Fisher Scientific, Waltham, United States). Obtained 16S rDNA sequences were compared with those available in the BIBI IV 16S Automated ProKaryotes Phylogeny database (https://lbbe.univ-lyon1.fr/fr). Sequence identity thresholds determined taxonomic assignment: ≥97-99% identity corresponded to genus level, and ≥99% identity corresponded to species level. When the database assigned multiple species within the same group with ≥99% identity, the bacterium was identified at the group level. Sequences showing less than 97% identity were considered inconclusive.

### Bioinformatic analyses

Reads with low quality scores were removed using Trim Galore software (10). A quality threshold of 30 (Phred score) was applied. The quality of the remaining reads was assessed using FastQC software (11). Reads originating from human DNA (hg38) were removed by mapping them to the human genome with Bowtie2 (12) Bacterial species were identified using MetaPhlAn4 (13). The results were then compared to those obtained through traditional 16S rDNA Sanger sequencing. Centrifuge2 (14). was used to quantify the proportion of reads originating from human DNA (remaining after mapping to the human genome). It was also used in conjunction with MetaPhlAn3 (15)) as secondary tools to investigate and explain any discrepancies observed between the two identification methods as second-line tools to explain discrepancies.

### Statistical analyses

Descriptive statistics (N, median (interquartile range) for quantitative and n (%) for qualitative) are presented in the Supplementary Table 1. Univariate analyses based on Wilcoxon rank-sum, Kruskal-Wallis, or Fisher tests (as appropriate depending on the quantitative/qualitative nature of examined variables and number of categories) were reported descriptively and not adjusted for multiplicity. For exploratory analysis of sample characteristics associated with diagnostic yield at the sample level, the following variables of interest were selected: (1) the 16S qPCR amplification cycle threshold (Ct), (2) the number of bacterial species found, (3) the number of reads assigned to the dominant non-human species, (4) the number of reads assigned to non-human species and (5) the number of reads assigned to the human genome. Variables 2-5 were obtained from Centrifuge2.

For multivariate analyses, read counts were expressed as log(reads+1)/log(2). We used univariate logistic regression to find associations between these variables and different clinical cases of interest, in different populations: 16S failure among all samples, CMg failure among samples with 16S failure, CMg-16S agreement among samples with a 16S diagnosis.

In order to build a multivariate model, a multiple-step approach was used, first to obtain predictors, then to control for false discovery rate, and finally to shrink coefficient estimates to avoid overfitting. First, all possible models based on a pre-specified list of potential predictors were compared based on AIC, and the best was chosen. Second, all variables with a Benjamini-Hochberg q-value > 0.1 were eliminated, in order to limit false discovery rate at 10%. Finally, an elastic net regression was run based on the R package glmnet (16) with default settings, and the penalization parameter was chosen so that the variables with FDR < 0.1 would have non-zero penalized coefficients.

Two sets of potential predictors were used in separate logistic regression models to predict the probability of an adjudicated CMg finding. The first set consisted of molecular biology variables to identify potential quality control indicators. These variables included: 16S PCR cycle threshold, number of reads mapped to major species (by Centrifuge), number of non-human reads (by Centrifuge), number of human reads (by Centrifuge), number of MetaPhlAn4 species, and maximum genome completeness (assessed by Quast) (17)), total amount of purified DNA, and number of library amplification rounds before sequencing (8 candidate variables total).

A second set of eight candidate variables was examined. These variables linked to the clinical situation and sample type were intended to inform pathologists on when to attempt CMg. They included: 16S PCR cycle threshold (again), heart valve sample type (yes/no), effusion fluid sample type (yes/no), abscess or vegetation (yes/no), antibiotic treatment before sampling (yes/no), suspected uncultivable or slow growth bacterium (yes/no), neutrophils on direct examination (yes/no), and bacteria on direct examination (yes/no). For these models, receiver operating characteristic (ROC) curves were generated. These curves plot sensitivity (ability to detect true CMg+ cases) against specificity (ability to correctly identify non-CMg+ cases) at different prediction probability thresholds.

Two ROC curves were created for each model: one using the original, unshrunk coefficient estimates, and another using the shrunk estimates obtained from the elastic net regression.

To assess the expected performance of these models on future samples, a leave-one-out cross-validation strategy was implemented. In this approach, a model was built on 72 samples, excluding one sample. The remaining sample was then used for prediction, and this process was repeated for all 73 samples. This resulted in 73 prediction probabilities for CMg+ results. By varying the prediction probability threshold, a final ROC curve (specificity vs. sensitivity) was derived. Again, this process was repeated using both the shrunk and unshrunk estimates.

All computations were performed using R version 4.2.2 (18).

## Results

### Population

In total, 50 patients were included and 73 samples were considered (mean of 1.4 samples per patient [1-4]). Clinical characteristics are detailed in Supplementary Table 1. The mean age of patients was 55 ± 17 years and 35/50 were male. Thirty-three out of fifty (66.0%) had at least one documented antibiotic initiation before sampling (Supplementary Table 2). The samples were cardiac valves (n=27, 17 patients), serous fluids (n=15, 13 patients), bone and joint samples (n=22, 11 patients), abscesses (n=6, 6 patients), lymph nodes (n=2, 2 patients) and scarpa sample (n=1, 1 patient).

### 16S rDNA Sanger sequencing results

Of the 73 samples analyzed, 30 (41.1%) from 23 patients tested positive for 16S rDNA Sanger sequencing. However, only 20 (27.4%) of these positive results, found in 17 patients, were considered clinically relevant. Bacteria found in 16S rDNA Sanger sequencing but not considered to be clinically relevant (n=10) were coagulase-negative staphylococci (n=8), *Bacillus* (n=1) and *Geobacillus* (n=1). Conversely, bacteria identified by 16S rDNA Sanger sequencing and considered to be clinically relevant (n=20) were *Staphylococcus aureus* (n=3), *Cutibacterium acnes* (n=3), *Streptococcus pneumoniae* (n=2), *Streptococcus oralis* (n=2), *Corynebacterium diphtheriae* (n=2), *Streptococcus intermedius* (n=1), *Streptococcus dysgalactiae* (n=1), *Streptococcus anginosus* (n=1), *Fusobacterium nucleatum* (n=1), *Francisella tularensis* (n=1), *Cutibacterium avidum* (n=1), *Bartonella henselae* (n=1) and *Aggregatibacter actinomycetemcomitans* (n=1) (Supplementary Table 3).

### Technical validation of the metagenomic pipeline

The sequencing technical results are detailed in the Supplementary Table 4. A total of 10 no-template controls (NTC) and 13 negative biological control (BC) were sequenced over 5 sequencing runs, with 5, 12 or 20 cycles applied during library preparation (Figure 1A). In NTC, MetaPhlAn4 detected 135 unique bacteria (40 over 1% abundance). Most abundant species included *Cutibacterium acnes* (mean abundance 35.4%), *Moraxella osloensis* (mean abundance 21.5%) and a bacterium identified as a species-level genome bin yet without clear taxonomy (GGB2722-SGB3663, mean abundance 11.1%, with SGB3663 being assigned as *Lawsonella cleavelandensis*). Of note when preparing NTC libraries with 12 cycles, only 2 NTC yielded bacteria: *C. acnes* (n=2), *M. osloensis* (n=2), *Ralstonia pickettii* (n=1) and *Actinomyces oris* (n=1). In BC (n=13), only samples with 12 or 20 cycles of library amplification yielded 20 unique bacteria, with 9 occurrences over 1% relative abundance. As for NTC, *C. acnes* was found with the highest abundance (Supplementary Tables 3 and 4). Interestingly, for a given number of amplification cycles NTC/BCs and clinical samples found negative in CMg had similar composition (Figure 1B).

**Figure 1:**
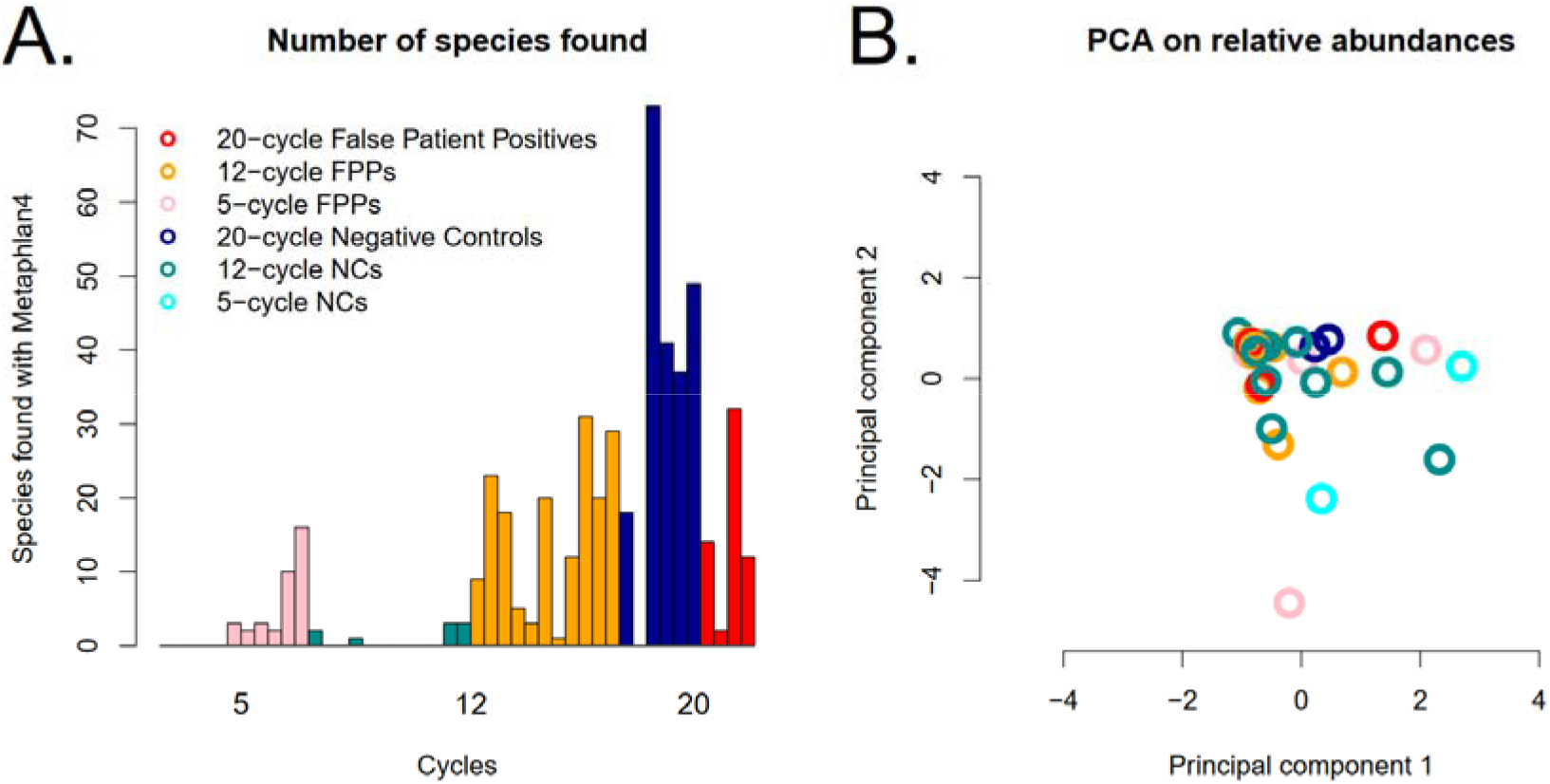
CMg number of species found (panel A) and principal component analysis (panel B) for the false patient positives and negative controls (with or without template, NTC and BC). Color codes are the same in the two panels.

### Clinical samples’ metagenomic sequencing results

In the clinical samples, we identified a total of 363 microorganisms (from 124 unique taxa, with 61 o1% abundance, Supplementary Table 5). At the sample level the sensitivity of CMg was 65.0% (13/20) compared to 16S rDNA Sanger sequencing as gold standard (Table 1). At the patient level, the sensitivity of CMg was 70.6% (12/17) (Table 2).

**Table 1:** Results from clinical metagenomics (CMg) and 16S rDNA Sanger sequencing when considering samples (n=73)

| Sample level |  | 16S rDNA Sanger sequencing |  |
| --- | --- | --- | --- |
|  |  | Positive | Negative |
| **CMg** | Positive | 15* | 10 |
|  | Negative | 5 | 43 |

\*Agreement on 13/15 samples

**Table 2:** Results from clinical metagenomics (CMg) and 16S rDNA Sanger sequencing when considering patients (n=50)

| Patient level |  | 16S rDNA Sanger sequencing |  |
| --- | --- | --- | --- |
|  |  | Positive | Negative |
| **CMg** | Positive | 14* | 8 |
|  | Negative | 3 | 25 |

\*Agreement on 12/14 patients

Discrepancies between 16S rDNA Sanger sequencing and CMg are detailed in Table 3. In seven samples, bacteria found by 16S rDNA Sanger were not detected by CMg. Among them, some bacteria not detected by MetaPhlAn4 were detected by MetaPhlAn3 or Centrifuge (n=5). In particular, MetaPhlAn4 did not detect a *Streptococcus anginosus,* while it detected other anaerobic bacteria. No clear explanation could be given for the 2 remaining samples.

**Table 3:** Details of the discrepancies observed between 16S rDNA Sanger sequencing (16S) and clinical metagenomics (CMg).

| Patient | Discrepancy | Number of samples | Culture (for this episode) | Details | Antibiotics |
| --- | --- | --- | --- | --- | --- |
| 23 | 16S+/CMg- | 4 | Negative | Relapse of a shoulder infection caused by *Cutibacterium acnes* (found in culture) despite 6 weeks of amoxicillin. *C. acnes* found in 16S (2/4) for the new episode, while culture was negative. | Amoxicillin+rifampicin, then levofloxacin+rifampicin |
| 27 | 16S+/CMg- | 1 | Negative | Tularemia, with arm abscess positive in 16S for *Francisella tularensis*, not detected by MetaPhlAn4 but detected by MetaPhlAn3. | Doxycycline |
| 31 | 16S+/CMg- |  | Negative | Drainage of a possibly infected deep haematoma of the scarpa, not in contact with the prosthesis. *Staphylococcus* spp (species close to *S. aureus*) found in 16S. | Cefepime+daptomycin, stopped before 16S results. |
| 1 | 16S-/CMg+ | 1 | Negative | Monoarthritis, knee synovial fluid with 100% reads assigned to *Streptococcus intermedius*. Patient treated for tuberculosis | None for this episode |
| 7 | 16S-/CMg+ | 2 | Negative | Post-infectious inflammation, 1/2 pericardic fluid with several bacteria (including anaerobic bacteria) found in CMg | Cefotaxime 2 days, stopped after negative cultures |
| 15 | 16S-/CMg+ | 3 | Negative | Ankle osteitis with material. 1/3 with 100% reads assigned to *Bacteroides thetaiotaomicron*. | Ceftriaxone+daptomycin, then ceftriaxone+linezolid (25 days, stopped after negative 16S) |
| 18 | 16S-/CMg+ | 1 | Negative | Ascitis in a patient treated for osteitis caused by *Pseudomonas aeruginosa*. CMg finds *Staphylococcus aureus*, *Enterococcus faecalis*, *Pseudomonas aeruginosa* and *Escherichia coli* | Ceftazidime+amikacin, then ceftazidime+ciprofloxacin |
| 36 | 16S-/CMg+ | 1 | Negative | Infective endocarditis caused by *Enterococcus faecalis* (blood culture). CMg finds *E. faecalis*. | Amoxicillin |
| 41 | 16S-/CMg+ | 1 | Negative | Drained pleuropneumopathy not documented after drainage and antibiotic therapy. CMg finds *Enterococcus faecium* and *Paracoccus haematequi*. | Levofloxacin, then aztreonam linezolid |
| 43 | 16S-/CMg+ | 2 | Negative | Infective endocarditis on native mitral and aortic valves with negative blood cultures, probably of dental origin, decapitated on Amoxicillin-Gentamicin. CMg finds *Streptococcus equinus* (1/2 samples). | Amoxicillin+gentamicin |
| 47 | 16S-/CMg+ | 1 | Negative | Septic arthritis on material in the right shoulder. CMg finds *Cutibacterium acnes* in a shoulder fluid. | Cefepime+daptomycin, then levofloxacin+clindamycin |

Among samples with a negative 16S rDNA Sanger sequencing, 31 had at least one identification with MetaPhlAn4 in CMg, with 349 bacteria found. In 21 samples, bacteria were deemed clinically irrelevant, of which 19 had *Cutibacterium acnes* as the dominant species. Among the other 10 samples (from 10 patients), the following species were found: *Streptococcus intermedius*, *Enterococcus faecalis*, *Enterococcus faecium*, *Paracoccus haematequi*, *Bacteroides vulgatus*, *Staphylococcus aureus* and *Streptococcus equinus*. In one patient, various anaerobic bacteria were found. Among these 10 patients, 3 had another sample for which 16S rDNA Sanger sequencing was positive, leaving seven patients for whom the microbiological diagnostic could only be provided by CMg (five infective endocarditis, one bone and joint infection and one peritonitis).

### Factors associated with 16S rDNA Sanger sequencing and CMg findings

We compared 16S qPCR results between patients with significant bacteria identified by 16S rDNA Sanger sequencing and CMg, and those with no bacteria detected. As expected, Ct values differed significantly between the two groups (Figure 2). Ct values above 23 resulted in positive or negative results for both 16S rDNA Sanger sequencing and CMg assays. In contrast, Ct values below 23 yielded positive results in all CMg runs and all but one 16S rDNA Sanger sequencing run. In multivariate analysis, the Ct value was the only consistent factor associated with a positive CMg diagnosis (univariate OR per additional cycle, 0.67, p=0.008) (Supplementary Table 6).

**Figure 2:**
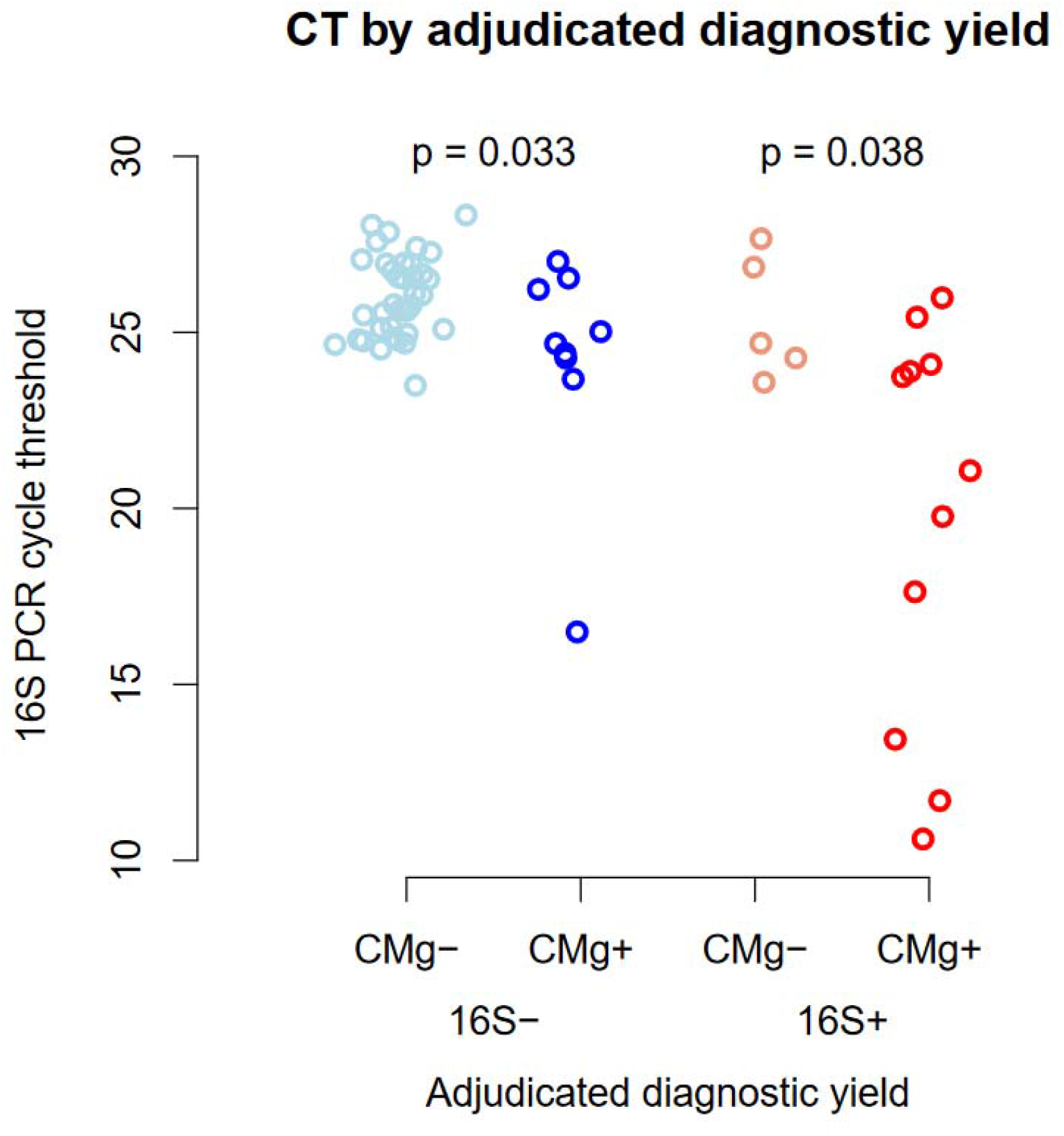
16S qPCR cycle threshold (Ct) values according to the identification of a clinically-significant bacterium by 16S rDNA Sanger sequencing (16S- or 16S+) or CMg (CMg- or CMg+). p-values by the rank sum test within 16S subgroups. Missing Ct values not shown.

When analyzing molecular biology data to predict CMg+ results, four factors were included in the final model with a 10% false discovery rate (Figure 3A): 16S qPCR Ct value, number of CMg species identified, number of non-human CMg reads and, highest genome completeness among CMg species (%). The derived unregularized score was -0.833 × (16S Ct - 24.18) + -0.055 × (Highest genome completeness (%) - 63.45) + -0.182 × (Number of species - 4.5) + 0.507 × (log(1+number of reads mapped to dominant species)/log(2) - 10.83); its derivation set AUC was 0.91, its LOCV AUC was 0.86; the threshold with 95% NPV on the LOCV set was -1.14 (below this value, chances for a CMg+ relevant finding were very low). For the regularized score, the formula was -0.227 × (16S Ct - 24.18) + -0.006 × (Highest genome completeness (%) - 63.45) + -0.016 × (Number of species - 4.5) + 0.067 × (log(1+number of reads mapped to dominant species)/log(2) - 10.83) + 0.026 × (log(1+number of reads mapped to bacteria)/log(2) - 11.33); its derivation set AUC was 0.88, its LOCV AUC was 0.75, and no threshold yielded 95% NPV.

**Figure 3:**
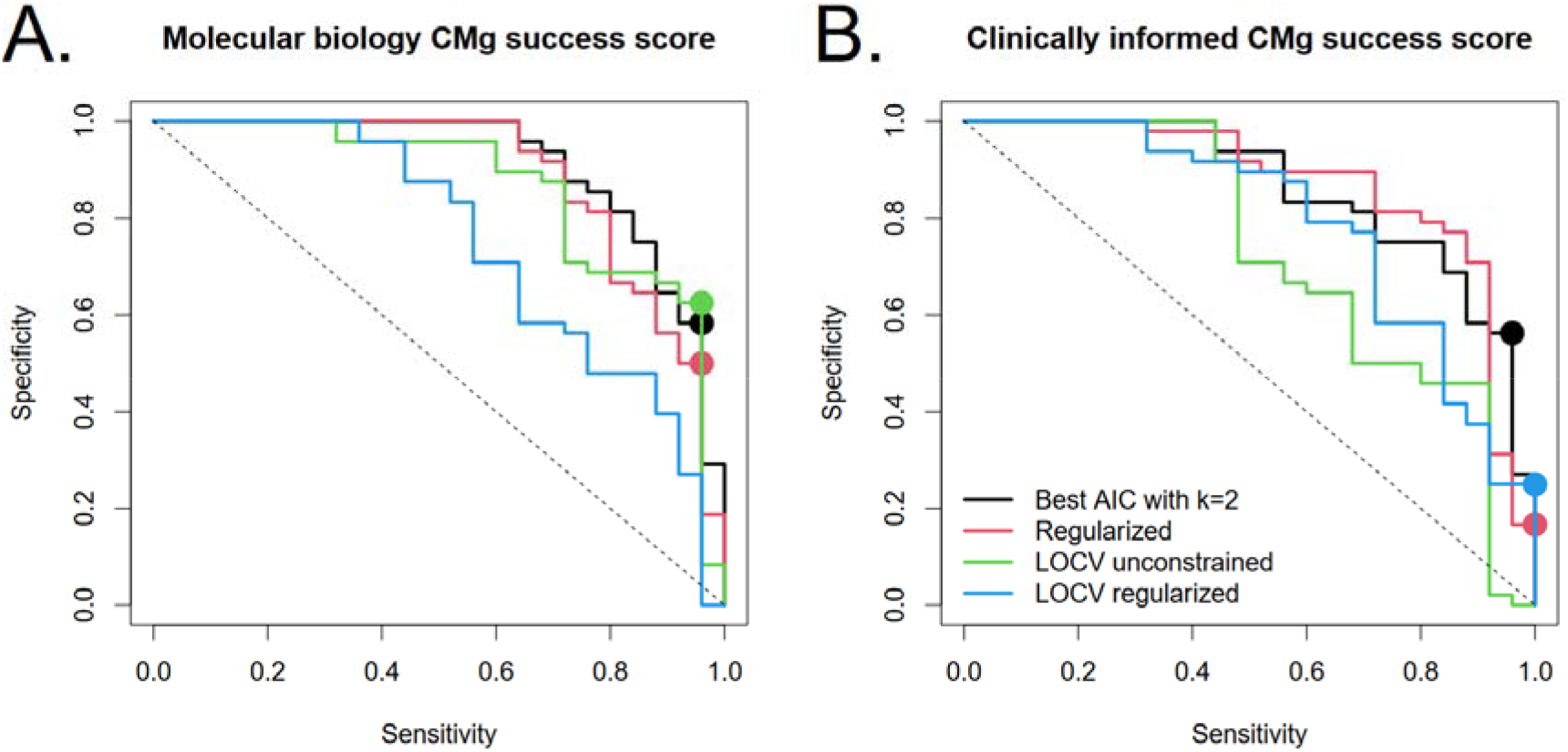
Prediction scores for positive CMg diagnostic yield and their receiver operating characteristic curves for the original and leave-one-out cross-validation (LOCV) datasets, with or without elastic net regularization. Dots indicate thresholds with observed NPV of 95% (on the relevant dataset). (A) Score derived from molecular biology variables (see Methods for initial set); the final model contains 16S cycle threshold (Ct), number of CMg species, number of CMg non-human reads, and CMg highest genome completeness Ct (B) Score derived from 16S Ct and clinical history variables (see Methods for initial set); the final model contains 16S Ct, heart valve sample type, and pre-treatment with antibiotics. Points represent thresholds with 95% negative predictive value (thus advising against attempting CMg), when they existed (see Results for thresholds and scores).

When trying to predict CMg+ results from non-CMg data (mostly clinical), 3 predictors were retained in the model with 10% false-discovery rate (Figure 3B): 16S qPCR Ct, heart valve sample type, and sequencing request because of pre-treatment with antibiotics. The derived unregularized score was - 1.024 × (16S Ct - 24.18) + 1.939 × (heart valve sample indicator) + 1.363 × (Pre-treatment with antibiotics indicator); its derivation set AUC was 0.86, its LOCV AUC was 0.73, and no threshold yielded 95% NPV on the LOCV set. For the regularized score, the formula was -0.200 × (16S Ct - 24.18) + 0.361 × (Heart valve sample indicator) + 0.034 × (Pre-treatment with antibiotics indicator); its derivation set AUC was 0.87, its LOCV AUC was 0.79, and the threshold with 95% NPV was -0.27 (below this value, chances for a CMg+ finding were very low).

### Metagenomic results beyond species identification

Among 32 bacteria identified in the 25 CMg+ samples, the mean genome completeness was 65.5% (range 14.0-98.4%), with 13 over 80% (Supplementary Table 7). Antibiotic resistance genes (ARG) were found in 17 samples (23.2%, 17/73), for a total of 15 distinct ARG and 37 occurrences (1 to 4 ARG per sample, Supplementary Table 8). In the NTC (n=10), 13 distinct ARG were found (21 occurrences, from 0 to 9 per sample), including mostly genes encoding for aminoglycosides-modifying enzymes (n=10), but also genes encoding for carbapenemases (*bla*_KPC_, n=2). Six ARG were found in both patients and NTC: *aph(3’)-IIa* (found in 8 samples), *ant(3’’)-Ia* (7 samples), *aph(3’)-Ia* (5 samples), *bla*_TEM-116_ (3 samples), *bla*_TEM-1C_ (1 sample) and *fosB_3* (1 sample).

Among bacteria with >80% completeness, 3 *S. aureus* were identified with no methicillin-resistance associated gene (e.g. *mecA*), while a penicillinase-encoding gene was detected (*blaZ*). Accordingly, the 3 strains could be considered as methicillin-susceptible. In sample 44, an *E. coli* was identified, with no ARG potentially associated with beta-lactam resistance in Enterobacterales.

### Comparison of 16S rDNA sequencing and CMg using other study results

In addition to the results of the present study, we considered the results we obtained in a previous work focusing on bone and joint infections (8) and in the studies by Lamoureux e*t al* (7) and Gu *et al* (9). In total, 242 samples from 156 patients could be analyzed (Table 4). The overall sensitivity of CMg when compared to 16S rDNA sequencing was 89.3% (67/75). Besides, CMg was positive (with bacteria not considered to be contaminants) in 39.7% (60/151) samples.

**Table 4:** Results from the present study and other studies (7–9) comparing 16S rDNA sequencing (referred to as “16S” in the table) and clinical metagenomics (“CMg” in the table).

| Study | Number of samples | Number of patients | Proportion of samples with 16S+ | Sensitivity (sample level) | Proportion of samples CMg+/16S- |
| --- | --- | --- | --- | --- | --- |
| This study | 73 | 50 | 27.4% (20/73) | 65.0% (13/20) | 18.9% (10/53) |
| d'Humières *et al.* 2022 | 94 | 34 | 30.8% (29/94) | 100.0% (29/29) | 62.7% (42/67) |
| Lamoureux *et al.* 2022 | 67 | 64 | 38.8% (26*/67) | 96.2% (25/26*) | 19.5% (8/41) |
| Gu *et al.* 2022 | 8 | 8 | 87.5% (7/8) | 75.0% (6/8) | 12.5% (1/8) |
| Total | 242 | 156 | 33.9% (82/242) | 88.0% (73/83) | 36.1% (61/169) |
| *after clinical adjudication |  |  |  |  |  |

## Discussion

This study compared the performance of CMg to the 16S rDNA Sanger sequencing method for diagnosing bacterial infections. CMg displayed a sensitivity of 65.0% (at the sample level) and 70.6% (at the patient level) compared to 16S rDNA Sanger sequencing. This sensitivity was lower than the sensitivity found in a previous study from our group (100%) and in Lamoureux *et al*. (7) (96.2%). For two samples, the non-detection of bacteria found in 16S rDNA Sanger sequencing can be explained by the bioinformatic pipeline based on MetaPhlAn which has a conservative approach in detriment to sensitivity (19). In another sample, CMg identified several anaerobic bacteria while 16S rDNA Sanger sequencing identified a *S. anginosus*. In this case, both methods could be considered as partially correct. We previously observed a similar situation in mandibular infection where culture only identified a *S. anginosus* while CMg identified 38 bacterial species (anaerobic bacteria) (20). This nicely shows the complementarity between conventional methods and CMg.

In addition, CMg identified bacteria missed by 16S rDNA Sanger sequencing in 10 samples, which is in the range of the previous findings by Lamoureux *et al*. (7) and Gu *et al*.(9), yet lower than in our previous study on bone and joint infections (8). A theoretical advantage of CMg over 16S rDNA Sanger sequencing is the capacity to detect multiple microorganisms in a sample while 16S rDNA is limited to a very few. Indeed, we had 2 samples negative for 16S rDNA sequencing and positive for more than 1 bacteria in CMg. Still, 8 samples positive in CMg and negative in 16S rDNA sequencing had only one bacterium found in CMg, suggesting that the capacity of CMg to detect additional bacteria over 16S rDNA Sanger sequencing does not only rely on its capacity to detect multiple bacteria.

Beyond the comparison between CMg and 16S rDNA Sanger sequencing, we looked for factors affecting the performances of the methods. As expected, 16S qPCR Ct values were associated with both CMg and 16S rDNA Sanger sequencing results in that lower Ct values correlated with a higher likelihood of positive results for both methods. However, we did not identify a value above which a test was always negative in CMg, ruling out the possibility of using a simple 16S qPCR as a first-line test supporting the performance of CMg. Nonetheless, the 16S qPCR value together with other clinical, biological and bioinformatic data could help in interpreting CMg data. In a previous work, Janes *et al.* similarly combined CMg results together with biological data (product of the sample’s DNA concentration with human fraction of reads and the pathogenicity of bacteria) to better interpret CMg results (21). Our observations support the approach that CMg should not be considered as a standalone test and that clinical, biological and bioinformatic data could be considered to help with the interpretation of CMg results. We also observed that the higher the number of amplification cycles (due to the low initial bacterial inoculum), the higher the number of contaminants. This raises the question of enriching each sample with exogenous non-human, non-microbial DNA to ensure sufficient DNA inoculum and thereby a minimal number of amplification cycles.

The study has limitations: the small sample size (73 samples from 50 patients, mostly from one site) might limit the generalizability of the findings. Besides, the predictive models require further validation on larger and more diverse datasets.

In conclusion, this study demonstrates the potential of CMg as a complementary tool for bacterial diagnosis, particularly for cases where 16S rDNA Sanger sequencing fails. CMg offers the advantage of identifying a wider range of bacteria, including uncultivable species and those resistant to antibiotics. However, further research is needed to optimize CMg protocols, establish robust reference standards, and validate predictive models for broader clinical application.

## Data Availability

All data produced in the present study are available upon reasonable request to the authors

## Acknowledgements

The authors are grateful to the staff of the Bichat and Beaujon hospitals who were involved in the inclusion of patients.

## Conflict of interest

All authors: none.

## Funding

This work was supported by a grant from the Assistance Publique–Hôpitaux de Paris (Contrat de Recherche Clinique).

## Notes

### Competing Interest Statement

The authors have declared no competing interest.

### Funding Statement

This work was supported by a grant from the Assistance Publique Hopitaux de Paris (Contrat de Recherche Clinique).

### Author Declarations

The project was approved by the Comite de Protection des Personnes Sud-Ouest et Outre Mer 1 on October 25, 2021 (2019-A01117-50 CPP 1-19-077 / SI 5294).

## References

1. Fida M, Khalil S, Abu Saleh O, Challener DW, Sohail MR, Yang JN, Pritt BS, Schuetz AN, Patel R. 2021. Diagnostic Value of 16S Ribosomal RNA Gene Polymerase Chain Reaction/Sanger Sequencing in Clinical Practice. Clinical Infectious Diseases 73:961– 968.

2. d’Humières C, Salmona M, Dellière S, Leo S, Rodriguez C, Angebault C, Alanio A, Fourati S, Lazarevic V, Woerther P-L, Schrenzel J, Ruppé E. 2021. The Potential Role of Clinical Metagenomics in Infectious Diseases: Therapeutic Perspectives. Drugs 81:1453–1466.

3. Chiu CY, Miller SA. 2019. Clinical metagenomics. Nat Rev Genet 20:341–355.

4. Abayasekara LM, Perera J, Chandrasekharan V, Gnanam VS, Udunuwara NA, Liyanage DS, Bulathsinhala NE, Adikary S, Aluthmuhandiram JVS, Thanaseelan CS, Tharmakulasingam DP, Karunakaran T, Ilango J. 2017. Detection of bacterial pathogens from clinical specimens using conventional microbial culture and 16S metagenomics: a comparative study. BMC Infect Dis 17:631.

5. Flurin L, Hemenway JJ, Fisher CR, Vaillant JJ, Azad M, Wolf MJ, Greenwood-Quaintance KE, Abdel MP, Patel R. 2022. Clinical Use of a 16S Ribosomal RNA Gene-Based Sanger and/or Next Generation Sequencing Assay to Test Preoperative Synovial Fluid for Periprosthetic Joint Infection Diagnosis. mBio 13:e0132222.

6. Mao Y-C, Chuang H-N, Shih C-H, Hsieh H-H, Jiang Y-H, Chiang L-C, Lin W-L, Hsiao T-H, Liu P-Y. 2021. An investigation of conventional microbial culture for the Naja atra bite wound, and the comparison between culture-based 16S Sanger sequencing and 16S metagenomics of the snake oropharyngeal bacterial microbiota. PLoS Negl Trop Dis 15:e0009331.

7. Lamoureux C, Surgers L, Fihman V, Gricourt G, Demontant V, Trawinski E, N’Debi M, Gomart C, Royer G, Launay N, Le Glaunec J-M, Wemmert C, La Martire G, Rossi G, Lepeule R, Pawlotsky J-M, Rodriguez C, Woerther P-L. 2022. Prospective Comparison Between Shotgun Metagenomics and Sanger Sequencing of the 16S rRNA Gene for the Etiological Diagnosis of Infections. Frontiers in Microbiology 13.

8. d’Humières C, Gaïa N, Gueye S, de Lastours V, Leflon-Guibout V, Maataoui N, Duprilot M, Lecronier M, Rousseau M-A, Gamany N, Lescure F-X, Senard O, Deconinck L, Dollat M, Isernia V, Le Hur A-C, Petitjean M, Nazimoudine A, Le Gac S, Chalal S, Ferreira S, Lazarevic V, Guigon G, Gervasi G, Armand-Lefèvre L, Schrenzel J, Ruppé E. 2022. Contribution of Clinical Metagenomics to the Diagnosis of Bone and Joint Infections. Frontiers in Microbiology 13:863777.

9. Gu W, Deng X, Lee M, Sucu YD, Arevalo S, Stryke D, Federman S, Gopez A, Reyes K, Zorn K, Sample H, Yu G, Ishpuniani G, Briggs B, Chow ED, Berger A, Wilson MR, Wang C, Hsu E, Miller S, DeRisi JL, Chiu CY. 2021. Rapid pathogen detection by metagenomic next-generation sequencing of infected body fluids. Nature Medicine 27:115–124.

10. 2012. Trim Galore.

11. 2015. FastQC.

12. Langmead B, Salzberg SL. 2012. Fast gapped-read alignment with Bowtie 2. Nature Methods 9:357–359.

13. Blanco-Míguez A, Beghini F, Cumbo F, McIver LJ, Thompson KN, Zolfo M, Manghi P, Dubois L, Huang KD, Thomas AM, Nickols WA, Piccinno G, Piperni E, Punčochář M, Valles-Colomer M, Tett A, Giordano F, Davies R, Wolf J, Berry SE, Spector TD, Franzosa EA, Pasolli E, Asnicar F, Huttenhower C, Segata N. 2023. Extending and improving metagenomic taxonomic profiling with uncharacterized species using MetaPhlAn 4. Nat Biotechnol 1–12.

14. Kim D, Song L, Breitwieser FP, Salzberg SL. 2016. Centrifuge: rapid and sensitive classification of metagenomic sequences. Genome Res 26:1721–1729.

15. Truong DT, Franzosa EA, Tickle TL, Scholz M, Weingart G, Pasolli E, Tett A, Huttenhower C, Segata N. 2015. MetaPhlAn2 for enhanced metagenomic taxonomic profiling. Nat Methods 12:902–903.

16. Friedman J, Hastie T, Tibshirani R, Narasimhan B, Tay K, Simon N, Qian J, Yang J. 2023. glmnet: Lasso and Elastic-Net Regularized Generalized Linear Models (4.1-8).

17. Gurevich A, Saveliev V, Vyahhi N, Tesler G. 2013. QUAST: quality assessment tool for genome assemblies. Bioinformatics 29:1072–1075.

18. R Development Core Team. R: A language and environment for statistical computing. (3.1.2). R Foundation for Statistical Computing, Vienna, Austria.

19. Sczyrba A, Hofmann P, Belmann P, Koslicki D, Janssen S, Dröge J, Gregor I, Majda S, Fiedler J, Dahms E, Bremges A, Fritz A, Garrido-Oter R, Jørgensen TS, Shapiro N, Blood PD, Gurevich A, Bai Y, Turaev D, DeMaere MZ, Chikhi R, Nagarajan N, Quince C, Meyer F, Balvočiūtė M, Hansen LH, Sørensen SJ, Chia BKH, Denis B, Froula JL, Wang Z, Egan R, Don Kang D, Cook JJ, Deltel C, Beckstette M, Lemaitre C, Peterlongo P, Rizk G, Lavenier D, Wu Y-W, Singer SW, Jain C, Strous M, Klingenberg H, Meinicke P, Barton MD, Lingner T, Lin H-H, Liao Y-C, Silva GGZ, Cuevas DA, Edwards RA, Saha S, Piro VC, Renard BY, Pop M, Klenk H-P, Göker M, Kyrpides NC, Woyke T, Vorholt JA, Schulze-Lefert P, Rubin EM, Darling AE, Rattei T, McHardy AC. 2017. Critical Assessment of Metagenome Interpretation-a benchmark of metagenomics software. Nature Methods 14:1063–1071.

20. Ruppé E, Lazarevic V, Girard M, Mouton W, Ferry T, Laurent F, Schrenzel J. 2017. Clinical metagenomics of bone and joint infections: a proof of concept study. Scientific Reports 7:7718.

21. Janes VA, Matamoros S, Munk P, Clausen PTLC, Koekkoek SM, Koster LAM, Jakobs ME, Wever B de, Visser CE, Aarestrup FM, Lund O, Jong MD de, Bossuyt PMM, Mende DR, Schultsz C. 2022. Metagenomic DNA sequencing for semi-quantitative pathogen detection from urine: a prospective, laboratory-based, proof-of-concept study. The Lancet Microbe 3:e588–e597.

